# Development of the Pruritus-Associated Stress Scale: A Cross-Sectional Pilot Study in Adults with Atopic Dermatitis, Chronic Prurigo and Chronic Pruritus on Non-Lesional Skin

**DOI:** 10.1101/2025.08.08.25333283

**Authors:** Svenja Royeck, Johanna Papathanassiou, Angelika Weigel, Nell Kindt, Bernd Löwe, Christian Mess, Claudia Zeidler, Felix Witte, Konstantin Agelopoulos, Henning Wiegmann, Stefan W. Schneider, Sonja Ständer

## Abstract

**Background:** A significant relationship exists between perceived stress and the exacerbation and perpetuation of chronic pruritic dermatoses. Despite this, there is a notable absence of validated tools to specifically measure pruritus-associated stress.

**Objective:** To develop and pilot the Pruritus-Associated Stress Scale (PASS), a patient-reported outcome measure (PROM) for assessing pruritus-associated stress.

**Patients and Methods:** Patients with chronic prurigo (CPG), atopic dermatitis (AD), and chronic pruritus on non-lesional skin (CPNL) were recruited at a German university centre. They were interviewed on pruritus-associated stress, and perceived stress using the PSS-10 and PSQ-30 questionnaires, to compile the first PASS version in accordance with the guidelines for PROM development. Subsequently, a second patient cohort was interviewed to refine the items of the PASS instrument based on impact analysis, interitem and item-total correlation, and internal consistency reliability.

**Results:** Of 55 patients (15 with AD, 20 with CPG, and 20 with CPNL; 61.8% female; mean age 61.0 ± 15.4 years), who participated in the item selection phase, 94.5% reported pruritus-associated stress in the previous two weeks. The preliminary PASS demonstrated excellent internal consistency (Cronbach’s alpha = 0.91). The twelve items that showed strong impact scores addressed nervousness, therapeutic strategies for managing pruritus-associated stress, fatigue, and urges to scratch more frequently or intensely due to pruritus.

**Conclusions:** This pilot study yielded a preliminary PASS, identified poorly performing items, and collected information for further refinement. As a next step, retaining the full item pool, an exploratory factor analysis will be conducted in a larger sample.

**SIGNIFICANCE:** This study addresses a critical gap in dermatological research by developing and piloting the first questionnaire specifically designed to assess pruritus-associated stress in patients with chronic pruritus of diverse aetiologies.

## INTRODUCTION

Chronic pruritus (CP) is “an unpleasant sensation of the skin and/or neighbouring mucous membranes commonly triggering an urge so scratch”, and can arise in the context of a wide variety of underlying diseases (1, 2). According to the classification by the International Forum for the Study of Itch (IFSI), CP can be classified into three distinct clinical categories: Pruritus occurring on lesional skin (group I), as seen in conditions such as atopic dermatitis (AD), pruritus on non-lesional skin (CPNL; group II) and pruritus associated with chronic scratch lesions (group III), such as in chronic prurigo (CPG) (3). There is growing recognition that CP is a persistent physical symptom (synonymous with a persistent somatic symptom). This implies that the pruritus can persist even after the resolution of the original trigger or underlying disease, thereby becoming a significant health burden in its own right (4, 5).

CP is frequently accompanied by marked impairment in quality of life (QoL), and many patients report elevated levels of anxiety and depression, sleep disturbances, stigmatization, body dysmorphic concerns, and reduced overall well-being, all symptoms related to stress (4, 5). In addition, perceived stress is recognized as an important modulator of CP in various dermatoses (6–8). Stress modulates the course of skin diseases through both neuroimmunological and behavioural pathways. A key example is the itch-scratch cycle, which can exacerbate CP and skin inflammation, ultimately comprising the skin’s barrier function (9–13). Thus, targeted stress-reducing interventions may beneficially modulate the course of CP (5). Addressing psychological comorbidities may contribute to improved pruritus outcomes (14).

However, a significant challenge in both research and clinical settings is the under-recognition of assessment and therapeutic approaches regarding pruritus-associated stress, a problem compounded by the lack of a validated instrument for its assessment. So far, stress has most frequently been assessed using generic self-reporting tools, such as the Perceived Stress Scale- 10 (PSS-10) or the Perceived Stress Questionnaire-30 (PSQ-30) (15, 16), which were not specifically designed for assessing pruritus-associated stress. Without a pruritus-specific instrument, it is neither possible to systematically investigate the relationship between stress and CP using biomarkers, nor can the effectiveness of pharmacological or non-pharmacological interventions targeting pruritus-associated stress be reliably determined.

Therefore, the primary objective of the present cross-sectional cohort study is to develop and pilot a new questionnaire specifically designed to measure pruritus-associated stress, the Pruritus-Associated Stress Scale (PASS). Moreover, we aim to investigate and compare perceived stress in patients with CP of different origins, i.e., AD, CPNL, and CPG.

## MATERIALS AND METHODS

### Study design and setting

This is a prospective cohort study that has been ongoing at the Center for Chronic Pruritus (KCP) in Münster, Germany. The methods used are in accordance with the current recommendations for the development of PROMs (16).

### Item generation

Following a narrative literature review to identify validated instruments for assessing perceived stress, a semi-structured interview was conducted with a cohort of adult CP patients (sufficiently fluent in the German language) who had AD, CPG, or CPNL (C1). Patients were asked a series of questions regarding pruritus-associated stress (**Tab. SI**), pruritus, pain, and scratching intensity, and requested to complete two validated stress questionnaires (PSS-10 and PSQ-30) (Figure 1).

**Figure 1:**
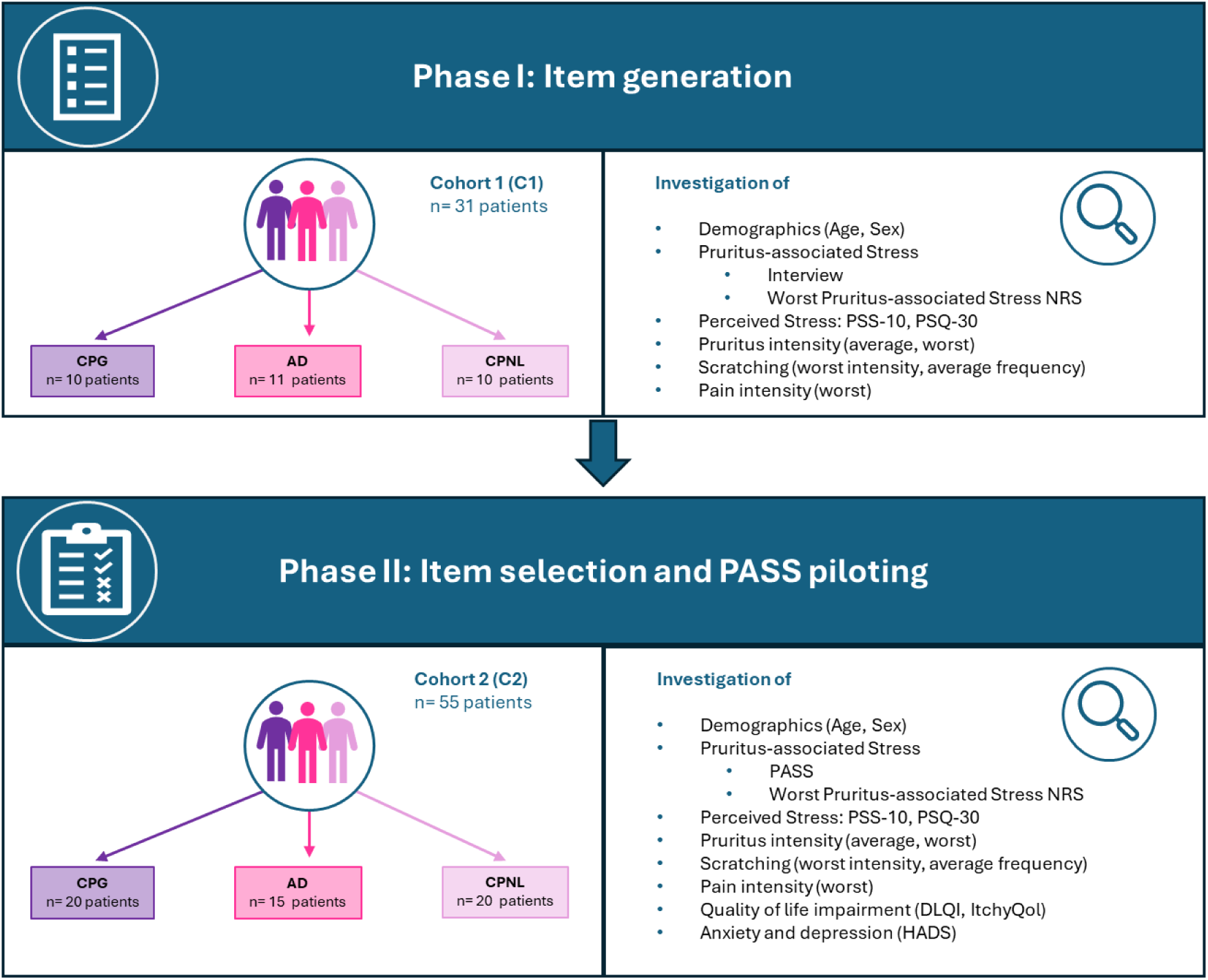
Study Flowchart. The figure presents both study phases (item generation and item selection) including recruited patient cohorts C1 and C2 as well as investigated demographics and patient-reported outcomes. AD: atopic dermatitis; CPG: chronic prurigo; CPNL: chronic pruritus on non-lesional skin; DLQI: Dermatology Life Quality Index; HADS: Hospital Anxiety and Depression Scale; PSS-10: Perceived Stress Scale; PSQ-30: Perceived Stress Questionnaire

Items from the interview were selected and incorporated into the preliminary PASS, if at least two-thirds of the patients had rated them as relevant (yes or no) for pruritus-associated stress. PSS-10 and PSQ-30 items were selected for inclusion in the preliminary PASS if they had an impact score in the upper third of distribution. The impact scores of the PSS-10 and PSQ-30 items were determined by first calculating the item importance: for each response option on the respective Likert scale, the respective scale value was multiplied by the number of patients who selected that option. These products were summed and then divided by the total number of responses to obtain the mean importance rating for each item. The item impact was then computed by multiplying the item importance by the proportion of patients who rated the item as important.

#### Item selection

A new analogous cohort of patients with CP (C2) was subsequently recruited and surveyed specifically regarding pruritus-associated stress using the preliminary PASS which was composed out of 25 items. Additional PROMs were used to evaluate perceived stress (PSS-10, PSQ-30) (15, 16); quality of life, including the Dermatology Life Quality Index (DLQI, range 0-30) (17) and the pruritus-specific ItchyQol (range 22-110) (18, 19); and anxiety and depression via the Hospital Anxiety and Depression Scale (HADS, ranges 0-21) (20) (**Figure 1**).

The pruritus intensity over the previous 24 hours was assessed using an NRS, capturing both average (AP-NRS) and maximum (WP-NRS) CP intensity (range 0–10) (21). Additionally, the average frequency of scratching, the highest intensity of scratching, and the highest intensity of pain experienced within the previous 24 hours were assessed using a NRS (0 = no, 10 = worst imaginable).

All participants provided both oral and written informed consent prior to enrolment. The study received approval from the Ethics Committee of the State Medical Association of Westfalen-Lippe, Münster, Germany (reference number: 2024-884-f-S).

### Interitem correlation and item-total correlation

Interitem and item-total correlations of the proposed PASS items were calculated to identify redundancies. To evaluate the internal structure of the preliminary PASS scale, bivariate Pearson’s inter-item correlations among all scale items were calculated. Values between 0.15 and 0.50 were considered optimal (22, 23). To assess the discriminative power of each item, corrected item-total correlations were calculated. Items with values below 0.30 may not adequately measure the underlying construct captured by the scale as a whole and were flagged for potential removal (23).

### Impact analysis

During the item selection phase, patients indicated on evaluation forms how often they had experienced the PASS items in the past two weeks (ranging from 0 to 4, with 0 indicating "never" and 4 indicating "very often") and rated the importance of each item (responding with "yes" or "no"). The frequency of each item was calculated as the percentage of respondents who had experienced the respective item. The item impact score was then derived by multiplying its frequency by the percentage of patients who had rated the item as important among all patients. Items with an impact score below 2.3 were considered of low relevance and might be excluded from the final version of the PASS.

### Internal consistency reliability

After the final set of items was determined through impact analysis, the internal consistency reliability of the scale was evaluated by calculating Cronbach’s alpha coefficient. The interpretation of Cronbach’s alpha values was as follows: below 0.60, unacceptable; 0.60 to 0.65, undesirable; 0.65 to 0.70, minimally acceptable; 0.70 to 0.80, respectable; and above 0.80, indicated very good internal consistency (23, 24).

#### Between-group comparisons analysis

Due to the exploratory study design, group comparisons of PROMs used the Kruskal-Wallis test without adjustments for multiple comparisons. If statistically significant differences were found between the three disease groups, pairwise comparisons between two groups were conducted using the Mann-Whitney U test.

##### Statistical analysis

All analyses were performed with SPSS software (IBM SPSS Statistics, version 29; IBM, Armonk, NY). Results with *p* < 0.05 were considered statistically significant.

## RESULTS

### Participants and patient-reported outcomes

Demographics and PROMs of all patients who participated in the item generation phase (C1; n = 31) and of all patients who took part in the item-selection phase (C2; n = 55) are presented in **Table I**.

**Table I:** Patient characteristics. AP-NRS: average pruritus of the previous 24 hours as assessed using a Numeric Rating Scale; DLQI: Dermatology Life Quality Index; HADS: Hospital Anxiety and Depression Scale; NRS: Numeric Rating Scale; PSQ-30: Perceived Stress Questionnaire; PSS-10: Perceived Stress Scale (PSS-10); PROMS: Patient Reported Outcome; SD: standard deviation; WPain-NRS: worst pain of the previous 24 hours as assessed using a Numeric Rating Scale WP-NRS: worst pruritus of the previous 24 hours as assessed using a Numeric Rating Scale

|  | Item-generation phase<br>(n= 31) | Item-selection phase<br>(n= 55) |
| --- | --- | --- |
| **Demographics** |  |  |
| Age (years), mean (SD) | 51.03 (15.64) | 61.02 (15.35) |
| Sex, n (%) |  |  |
| • Male, n (%) | 15 (48.4) | 20 (36.4) |
| • Female, n (%) | 16 (51.6) | 34 (61.8) |
| **Disease characteristics** |  |  |
| Chronic Prurigo, n (%) | 10 (32.3) | 20 (36.4) |
| Atopic Dermatitis, n (%) | 11 (35.5) | 15 (27.3) |
| Chronic Pruritus on non-lesional skin, n (%) | 10 (32.3) | 20 (36.4) |
| **PROMS** |  |  |
| PSS-10, mean (SD) | 17.84 (7.32) | 20.81 (7.23) |
| PSQ-30, mean (SD) | 0.41 (0.19) | 0.48 (0.20) |
| WP-NRS, mean (SD) | 2.39 (1.17) | 6.38 (2.28) |
| AP-NRS, mean (SD) | 2.03 (1.08) | 5.51 (2.20) |
| Worst Pruritus-associated Stress-NRS, mean (SD) | 5.20 (3.32) | 5.20 (2.72) |
| Worst Scratch Intensity-NRS, mean (SD) | 5.40 (2.96) | 5.51 (2.73) |
| Average Scratch Frequency-NRS, mean (SD) | 4.97 (2.90) | 5.24 (2.67) |
| WPain-NRS, mean (SD) | 3.84 (3.11) | 4.22 (3.21) |
| DLQI, mean (SD) | n.a. | 11.11 (7.84) |
| ItchyQol, mean (SD) | n.a. | 98.31 (36.03) |
| HADS – subscale anxiety, mean (SD) | n.a. | 8.76 (4.34) |
| HADS – subscale depression, mean (SD) | n.a. | 7.41 (3.96) |

A total of 55 patients participated in the item-selection phase (mean age 61.02 ± 15.35 years; 61.8% female; n = 15 with AD, n = 20 with CPG, and n = 20 with CPNL). The highest pruritus intensity during the past 24 hours was moderate (mean WP-NRS 6.38 ± 2.28), and the highest pain intensity was mild (mean worst-pain NRS 4.22 ± 3.21).

Quality of life was moderately to severely impaired (DLQI 11.11 ± 7.84; ItchyQoL 98.31 ± 36.03). Anxiety levels were elevated (8.76 ± 4.34) and depression levels were within normal ranges (**Table I**). Perceived stress was moderate (PSS-10 20.81 ± 7.23; PSQ-Index 0.48 ± 0.20).

#### PASS item selection

A total of 25 items from the questionnaire were analysed to assess reliability and item characteristics. The internal consistency of the scale was excellent, as indicated by a Cronbach’s alpha of 0.905 and a standardized Cronbach’s alpha of 0.910.

### Scale and summarized item statistics

Across all items, the means ranged from 1.60 (± 0.92) to 2.96 (± 1.11) (overall mean 2.38, range 1.36, max/min ratio 1.85, variance 0.19). On the scale level, the mean total score was 59.42 with a standard deviation of 13.75 and a variance of 189.14. Detailed item statistics (mean, standard deviation) are presented in **Table II**.

**Table II:** Item Descriptive and Item-Total Statistics. Only item 20 (highlighted in gray) exhibited a corrected item-total correlation value below 0.30 and is therefore considered for deletion from the final version of the PASS. This suggests that the item may not adequately measure the underlying construct assessed by the PASS as a whole.

| Item | Mean | SD | Item-Total Corr. | Cronbach's $\alpha$ if Item Deleted |
| --- | --- | --- | --- | --- |
| **Item 1:** How often did you feel nervous due to stress induced by pruritus in the last two weeks? | 2.87 | 0.90 | 0.56 | 0.90 |
| **Item 2:** How often did you feel restless due to stress induced by pruritus in the past two weeks? | 2.87 | 0.92 | 0.60 | 0.90 |
| **Item 3:** How often did you feel irritable due to stress induced by pruritus in the past two weeks? | 2.65 | 0.99 | 0.52 | 0.90 |
| **Item 4:** How often did you feel more prone to conflict due to stress induced by pruritus in the past two weeks? | 2.13 | 0.92 | 0.47 | 0.90 |
| **Item 5:** How often did your pruritus become stronger or more frequent in stressful situations in the past two weeks? | 2.51 | 1.09 | 0.64 | 0.90 |
| **Item 6:** How often did you feel the urge to scratch more frequently and/or more intensely in the past two weeks? | 2.95 | 0.93 | 0.42 | 0.90 |
| **Item 7:** How often did you try to distract yourself in order to cope with stress induced by pruritus in the past two weeks (e.g., through social contacts, hobbies, or relaxation exercises)? | 2.40 | 1.18 | 0.36 | 0.91 |
| **Item 8:** How often did you use therapeutic measures (e.g., application of cold, moisturizing the skin) to cope with stress induced by pruritus in the past two weeks? | 2.96 | 1.11 | 0.46 | 0.90 |
| **Item 9:** How often did you feel satisfied with your daily work and tasks and felt that you were doing something meaningful in the past two weeks? | 1.65 | 0.87 | 0.42 | 0.90 |
| **Item 10:** How often did you feel in good physical condition in the past two weeks? | 2.00 | 1.00 | 0.60 | 0.90 |
| **Item 11:** How often did you have difficulty relaxing in the past two weeks? | 2.44 | 1.03 | 0.39 | 0.90 |
| **Item 12:** How often did you feel burdened by experiences, conflicts, or arguments with other people in the past two weeks? | 2.18 | 1.11 | 0.48 | 0.90 |
| **Item 13:** How often did you have thoughts or worries that you could not suppress or could suppress only with difficulty in the past two weeks? | 2.29 | 0.94 | 0.50 | 0.90 |
| **Item 14:** How often did you feel confident that you were able to cope with your personal problems in the past two weeks? | 1.60 | 0.92 | 0.59 | 0.90 |
| **Item 15:** How often did you feel that you had everything under control in the past two weeks? | 2.02 | 0.99 | 0.64 | 0.90 |
| **Item 16:** How often did you feel annoyed by things over which you had no control in the past two weeks? | 2.35 | 0.99 | 0.43 | 0.90 |
| **Item 17:** How often did you feel tired in the past two weeks? | 2.96 | 1.00 | 0.37 | 0.90 |
| **Item 18:** How often did you feel that you were doing things you really enjoy in the past two weeks? | 1.91 | 0.89 | 0.59 | 0.90 |
| **Item 19:** How often did you have fun in the past two weeks? | 2.02 | 0.89 | 0.59 | 0.90 |
| **Item 20:** How often did you feel that you had too much to do in the past two weeks? | 2.45 | 1.27 | 0.11 | 0.91 |
| **Item 21:** How often did you feel safe and protected in the past two weeks? | 1.64 | 0.91 | 0.41 | 0.90 |
| **Item 22:** How often did you feel mentally exhausted in the past two weeks? | 2.87 | 1.00 | 0.68 | 0.90 |
| **Item 23:** How often did you feel tense in the past two weeks? | 2.82 | 0.91 | 0.75 | 0.90 |
| **Item 24:** How often did you feel rushed in the past two weeks? | 2.40 | 1.13 | 0.60 | 0.90 |
| **Item 25:** How often did you feel rested in the past two weeks? | 2.47 | 0.88 | 0.59 | 0.90 |

### Item-total statistics

Only item 20 exhibited an item-total correlation below 0.30, indicating that its removal from the final PASS should be considered in future evaluations. The remaining item-total correlations ranged between 0.36 and 0.75 (**Table II**).

### Inter-item correlations

The inter-item correlation matrix is presented in **Table III**. There were only a few inter-item correlations with coefficients of 0.7 or higher, which will need to be evaluated for potential removal in the next stage of the study.

**Table III:** Inter-item correlation matrix. Values present the Pearson’s correlation coefficient. Correlation coefficients ≥0.7 indicate extreme similarity and provide little additional informational value (redundancy; marked grey).

| Item | 1 | 2 | 3 | 4 | 5 | 6 | 7 | 8 | 9 | 10 | 11 | 12 | 13 | 14 | 15 | 16 | 17 | 18 | 19 | 20 | 21 | 22 | 23 | 24 | 25 |
| --- | --- | --- | --- | --- | --- | --- | --- | --- | --- | --- | --- | --- | --- | --- | --- | --- | --- | --- | --- | --- | --- | --- | --- | --- | --- |
| 1 | 1.00 | 0.85 | 0.66 | 0.40 | 0.26 | 0.52 | 0.36 | 0.39 | 0.35 | 0.33 | 0.18 | 0.17 | 0.29 | 0.27 | 0.29 | 0.26 | 0.04 | 0.38 | 0.26 | -0.05 | 0.03 | 0.25 | 0.61 | 0.23 | 0.40 |
| 2 | 0.85 | 1.00 | 0.72 | 0.35 | 0.31 | 0.59 | 0.40 | 0.50 | 0.36 | 0.40 | 0.22 | 0.22 | 0.26 | 0.33 | 0.33 | 0.21 | 0.04 | 0.42 | 0.25 | -0.12 | 0.10 | 0.22 | 0.53 | 0.23 | 0.42 |
| 3 | 0.66 | 0.72 | 1.00 | 0.60 | 0.19 | 0.42 | 0.38 | 0.62 | 0.08 | 0.32 | 0.26 | 0.16 | 0.25 | 0.32 | 0.27 | 0.34 | 0.04 | 0.32 | 0.28 | -0.23 | 0.25 | 0.18 | 0.41 | 0.13 | 0.21 |
| 4 | 0.40 | 0.35 | 0.60 | 1.00 | 0.27 | 0.18 | 0.28 | 0.40 | 0.10 | 0.22 | 0.15 | 0.25 | 0.13 | 0.15 | 0.24 | 0.50 | 0.09 | 0.26 | 0.29 | 0.06 | 0.25 | 0.28 | 0.38 | 0.38 | 0.06 |
| 5 | 0.26 | 0.31 | 0.19 | 0.27 | 1.00 | 0.30 | 0.29 | 0.22 | 0.39 | 0.41 | 0.31 | 0.38 | 0.38 | 0.51 | 0.40 | 0.46 | 0.12 | 0.43 | 0.39 | 0.26 | 0.47 | 0.44 | 0.44 | 0.52 | 0.38 |
| 6 | 0.52 | 0.59 | 0.42 | 0.18 | 0.30 | 1.00 | 0.09 | 0.29 | 0.16 | 0.20 | 0.10 | 0.10 | 0.32 | 0.21 | 0.20 | 0.22 | 0.18 | 0.24 | 0.18 | 0.07 | -0.05 | 0.21 | 0.30 | 0.34 | 0.33 |
| 7 | 0.36 | 0.40 | 0.38 | 0.28 | 0.29 | 0.09 | 1.00 | 0.40 | 0.17 | 0.13 | 0.36 | 0.20 | 0.09 | 0.20 | 0.28 | 0.18 | -0.07 | 0.12 | 0.15 | 0.03 | 0.09 | 0.22 | 0.24 | 0.10 | 0.17 |
| 8 | 0.39 | 0.50 | 0.62 | 0.40 | 0.22 | 0.29 | 0.40 | 1.00 | 0.20 | 0.24 | 0.34 | 0.25 | 0.26 | 0.43 | 0.31 | 0.10 | 0.17 | 0.20 | 0.23 | -0.19 | 0.21 | 0.31 | 0.27 | 0.06 | 0.21 |
| 9 | 0.35 | 0.36 | 0.08 | 0.10 | 0.30 | 0.16 | 0.17 | 0.20 | 1.00 | 0.41 | 0.26 | 0.26 | 0.29 | 0.41 | 0.38 | 0.08 | 0.01 | 0.37 | 0.27 | -0.09 | 0.10 | 0.33 | 0.42 | 0.11 | 0.41 |
| 10 | 0.33 | 0.40 | 0.32 | 0.22 | 0.39 | 0.20 | 0.13 | 0.24 | 0.41 | 1.00 | 0.05 | 0.17 | 0.34 | 0.57 | 0.82 | 0.21 | 0.22 | 0.50 | 0.54 | 0.06 | 0.39 | 0.37 | 0.51 | 0.39 | 0.38 |
| 11 | 0.18 | 0.22 | 0.26 | 0.15 | 0.31 | 0.10 | 0.36 | 0.34 | 0.26 | 0.05 | 1.00 | 0.32 | 0.19 | 0.05 | 0.10 | 0.25 | 0.29 | 0.33 | 0.29 | -0.10 | 0.17 | 0.36 | 0.34 | 0.17 | 0.28 |
| 12 | 0.17 | 0.22 | 0.16 | 0.25 | 0.38 | 0.10 | 0.20 | 0.25 | 0.26 | 0.17 | 0.32 | 1.00 | 0.48 | 0.18 | 0.22 | 0.49 | 0.22 | 0.22 | 0.22 | 0.16 | 0.31 | 0.37 | 0.35 | 0.38 | 0.29 |
| 13 | 0.29 | 0.26 | 0.25 | 0.13 | 0.38 | 0.32 | 0.09 | 0.26 | 0.29 | 0.34 | 0.19 | 0.48 | 1.00 | 0.40 | 0.31 | 0.27 | 0.33 | 0.21 | 0.33 | 0.07 | 0.28 | 0.32 | 0.37 | 0.26 | 0.30 |
| 14 | 0.27 | 0.33 | 0.32 | 0.15 | 0.51 | 0.21 | 0.20 | 0.43 | 0.41 | 0.57 | 0.05 | 0.18 | 0.40 | 1.00 | 0.58 | 0.20 | 0.29 | 0.28 | 0.40 | 0.06 | 0.38 | 0.45 | 0.45 | 0.34 | 0.47 |
| 15 | 0.29 | 0.33 | 0.27 | 0.24 | 0.40 | 0.20 | 0.28 | 0.31 | 0.38 | 0.82 | 0.10 | 0.22 | 0.31 | 0.58 | 1.00 | 0.15 | 0.34 | 0.49 | 0.50 | 0.11 | 0.42 | 0.49 | 0.52 | 0.42 | 0.39 |
| 16 | 0.26 | 0.21 | 0.34 | 0.50 | 0.46 | 0.22 | 0.18 | 0.10 | 0.08 | 0.21 | 0.25 | 0.49 | 0.27 | 0.20 | 0.15 | 1.00 | 0.11 | 0.14 | 0.14 | 0.09 | 0.25 | 0.37 | 0.30 | 0.36 | 0.13 |
| 17 | 0.04 | 0.04 | 0.04 | 0.09 | 0.12 | 0.18 | -0.07 | 0.17 | 0.01 | 0.22 | 0.29 | 0.22 | 0.33 | 0.29 | 0.34 | 0.11 | 1.00 | 0.23 | 0.40 | 0.06 | 0.11 | 0.63 | 0.42 | 0.39 | 0.42 |
| 18 | 0.38 | 0.42 | 0.32 | 0.26 | 0.43 | 0.24 | 0.12 | 0.20 | 0.37 | 0.50 | 0.33 | 0.22 | 0.21 | 0.27 | 0.49 | 0.14 | 0.23 | 1.00 | 0.63 | 0.02 | 0.33 | 0.53 | 0.60 | 0.37 | 0.46 |
| 19 | 0.26 | 0.25 | 0.28 | 0.29 | 0.39 | 0.18 | 0.15 | 0.23 | 0.27 | 0.54 | 0.29 | 0.22 | 0.33 | 0.40 | 0.50 | 0.14 | 0.40 | 0.63 | 1.00 | 0.11 | 0.46 | 0.48 | 0.42 | 0.43 | 0.27 |
| 20 | -0.05 | -0.12 | -0.23 | 0.06 | 0.26 | 0.07 | 0.03 | -0.19 | -0.09 | 0.06 | -0.10 | 0.16 | 0.07 | 0.06 | 0.11 | 0.09 | 0.06 | 0.02 | 0.11 | 1.00 | 0.08 | 0.22 | 0.14 | 0.57 | 0.09 |
| 21 | 0.03 | 0.10 | 0.25 | 0.25 | 0.47 | -0.05 | 0.86 | 0.21 | 0.10 | 0.39 | 0.17 | 0.31 | 0.28 | 0.38 | 0.42 | 0.25 | 0.11 | 0.33 | 0.46 | 0.08 | 1.00 | 0.23 | 0.19 | 0.31 | 0.13 |
| 22 | 0.25 | 0.22 | 0.18 | 0.28 | 0.44 | 0.21 | 0.22 | 0.31 | 0.33 | 0.37 | 0.36 | 0.37 | 0.32 | 0.45 | 0.49 | 0.37 | 0.63 | 0.53 | 0.48 | 0.22 | 0.23 | 1.00 | 0.67 | 0.54 | 0.62 |
| 23 | 0.61 | 0.53 | 0.41 | 0.38 | 0.44 | 0.30 | 0.24 | 0.27 | 0.42 | 0.51 | 0.34 | 0.35 | 0.37 | 0.45 | 0.52 | 0.30 | 0.42 | 0.60 | 0.42 | 0.14 | 0.19 | 0.67 | 1.00 | 0.51 | 0.67 |
| 24 | 0.23 | 0.23 | 0.13 | 0.38 | 0.52 | 0.34 | 0.10 | 0.06 | 0.11 | 0.39 | 0.17 | 0.38 | 0.26 | 0.34 | 0.42 | 0.36 | 0.39 | 0.37 | 0.43 | 0.57 | 0.31 | 0.54 | 0.51 | 1.00 | 0.44 |
| 25 | 0.40 | 0.42 | 0.21 | 0.06 | 0.38 | 0.33 | 0.17 | 0.21 | 0.41 | 0.38 | 0.28 | 0.29 | 0.30 | 0.47 | 0.39 | 0.13 | 0.42 | 0.46 | 0.27 | 0.09 | 0.13 | 0.62 | 0.67 | 0.44 | 1.00 |

### Item impact

As shown in **Table IV**, the range of calculated item impacts was between 1.46 and 2.96. Twelve out of 25 items achieved an item impact of 2.3 or higher and should therefore be considered for inclusion in the final version of the PASS. The three items with the highest item impact addressed therapeutic strategies for managing pruritus-associated stress, fatigue, and urges to scratch more frequently or intensely.

**Table IV:** Item Impact Scores and inter-item correlations. Questions considered suitable for the PASS according to an impact score ≥2.3 are marked yellow. For the final item selection, the inter-item correlations of items 1-3 must be considered.

| Items | Item content | Impact Score | Inter-item correlation ($r \geq 0.7$) |
| --- | --- | --- | --- |
| ***Feelings*** |  |  |  |
| 1 | Nervousness | 2.82 | Item 2 |
| 2 | Restlessness | 2.73 | Items 1, 3 |
| 3 | Irritability | 2.45 | Items 2 |
| 4 | Conflict proneness | 1.85 |  |
| 9 | Satisfaction with daily work | 2.23 |  |
| 10 | Good physical state | 1.90 | Item 15 |
| 12 | Strain from social contacts | 2.03 |  |
| 13 | Thoughts or worries | 2.04 |  |
| 14 | Feeling of problem-solving ability | 2.30 |  |
| 15 | Feeling of control | 1.88 | Item 10 |
| 16 | Anger over uncontrollable things | 2.09 |  |
| 20 | Feeling of being overwhelmed | 2.33 |  |
| 21 | Sense of safety | 2.20 |  |
| 22 | Mental exhaustion | 2.76 |  |
| 23 | Emotional tension | 2.68 |  |
| 24 | Feeling of being rushed | 2.28 |  |
| 25 | Feeling rested | 1.47 |  |
| ***Signs and symptoms*** |  |  |  |
| 5 | Pruritus (Intensity or frequency) | 2.41 |  |
| 17 | Fatigue | 2.85 |  |
| ***Coping and Therapies*** |  |  |  |
| 7 | Distraction | 2.26 |  |
| 8 | Therapeutic interventions | 2.96 |  |
| 11 | Relaxation | 2.31 |  |
| 18 | Valued activities | 1.86 |  |
| 19 | Fun | 1.90 |  |
| *Scratching* |  |  |  |
| 6 | Urge to scratch (frequency, intensity) | 2.83 |  |

#### Comparison of patients with AD, CPNL and CPG

The Kruskal-Wallis test revealed no statistically significant differences among the three disease groups with respect to age, gender, mean pruritus intensity over the past 24 hours, mean scratching frequency in the past 24 hours, highest scratching intensity in the last 24 hours, highest pain intensity in the last 24 hours, or anxiety and depression levels (all *p* > 0.05). There were also no significant differences in perceived stress levels (PSS-10: Kruskal-Wallis H(2) = 2.10, *p* = 0.35; PSQ-30: Kruskal-Wallis H(2) = 0.58, *p* = 0.75).

However, the groups differed significantly in terms of the worst pruritus intensity experienced during the last 24 hours (Kruskal-Wallis H(2) = 6.984, *p* = 0.030). Patients with CPNL reported a significantly higher worst pruritus intensity during the previous 24 hours than patients with AD (7.45 ± 2.09 vs. 5.40 ± 2.44, Mann-Whitney U = 77.00, Z = -2.46, *p* = 0.014).

Significant group differences emerged for three of the 25 potential PASS items: nervousness (item 1, Kruskal-Wallis H(2) = 6.08, *p* = 0.048), irritability (item 3, Kruskal-Wallis H(2) = 8.62, *p* = 0.013), and attempts to distract oneself to cope with pruritus-associated stress (item 7, Kruskal-Wallis H(2) = 7.63, *p* = 0.022). Specifically, patients with CPNL reported feeling nervous as a result of pruritus-associated stress significantly more frequently within the past two weeks than patients with AD (Mann-Whitney U = 87.50, Z = -2.19, *p* = 0.036). Patients with AD reported experiencing irritability due to pruritus-associated stress significantly more often than those with CPNL (Mann-Whitney U = 71.50, Z = -2.76, *p* = 0.008), and also patients with CPG reported irritability more frequently than CPNL patients (Mann-Whitney U = 125.50, Z = -2.13, *p* = 0.043). Regarding coping behaviours, patients with CPG reported significantly more frequent attempts to distract themselves in response to pruritus-associated stress compared to CPNL patients (Mann-Whitney U = 113.00, Z = -2.42, *p* = 0.018); similarly, such attempts were significantly more common among patients with AD than among those with CPNL (Mann-Whitney U = 85.50, Z = -2.25, *p* = 0.030).

## DISCUSSION

In CP, a complex, multidirectional interplay exists between pruritus-related stress, neuroimmune inflammation, and skin barrier dysfunction. Pruritus can induce stress and be induced by stress. Within this dynamic, stress also compromises the integrity of the skin barrier, rendering it more susceptible to further irritation and inflammation (25, 26). Therefore, developing a scale to specifically evaluate pruritus-associated stress is crucial for guiding appropriate treatment, not only to improve patients’ quality of life but also to meaningfully influence and modulate disease progression. Consequently, in this study, we included patients from all three clinical groups affected by CP: Patients with atopic dermatitis (IFSI I), chronic pruritus on non-lesional skin (IFSI III) and chronic prurigo (IFSI III).

Demographics were comparable among the three groups. The majority of assessed PROMs including pruritus and pain intensities as well as depression scores indicated moderate levels. In contrast, quality of life measures revealed severe impairment, while anxiety scores were predominantly within the borderline abnormal range. Interestingly, the perceived stress levels were also moderate, albeit higher than in the general population without CP (16, 27). Thus, the sample was suitable for the study’s objective of developing and piloting the PASS. The groups differed significantly in terms of the worst pruritus intensity experienced during the last 24 hours and ItchyQol. This was expected and welcome for the development of a PASS suitable for CP of various origins. It is known that these pruritus-affected groups differ in terms of scratching behaviour (CPNL: low-intensity, CPG: high-intensity), the high prevalence of CPNL, and the higher levels of the two most frequent comorbidities of CP (anxiety and depression) in CPNL and CPG compared to AD (28, 29).

The literature review, the structured interview, and the responses to the PSS-10 and PSQ-30 enabled the generation of 25 PASS items. The two questionnaires were included because they are validated and widely used for stress assessment, and we wanted to include suitable stress indicators in the PASS. Additionally, the PSS-10 was selected because there are existing studies in which perceived stress in patients with dermatological conditions was specifically measured using the PSS-10, providing relevant prior data for this population (30–35).

As part of this study, all 25 preliminary items were administered to assess their psychometric properties prior to item reduction and exploratory factor analysis (EFA) in a larger sample. The current analyses aimed to identify poorly performing items and inform item refinement, while retaining the full item pool for subsequent factor-analytic procedures.

Item-level statistics included descriptive indices (mean, standard deviation, skewness, and kurtosis), corrected item–total correlations, inter-item correlations, and a participant-weighted item impact score (i.e., importance × frequency). Items 1 (nervousness), 2 (restlessness), 3 (irritability), 5 (pruritus), 6 (urge to scratch), 8 (therapeutic interventions), 10 (good physical state), 18 (valued activities), and 22-25 (mental exhaustion, emotional tension, feeling of being rushed, feeling rested) showed satisfactory strong results across all indices and are considered psychometrically and substantively robust. Several items (e.g., 7 [distraction], 11-16 [relaxation, strain from social contacts, thoughts of worries, feeling of problem-solving ability, feeling of control, anger over uncontrollable things], 19 [fun]) demonstrated moderate performances, with acceptable but somewhat lower item-total correlations or limited relevance, suggesting the need for potential revision (Table IV).

Items 17 (fatigue) and 20 (feeling of being overwhelmed) proved to be problematic. Item 20 showed a very weak item-total correlation (rit = 0.11), numerous negative inter-item correlations, and limited experienced relevance, marking it as a likely candidate for removal. Item 17 was perceived as relevant (impact = 2.73) but showed a minimal correlation with the rest of the scale (mean r = 0.15), suggesting it may reflect a distinct construct or an interpretation issue.

Despite these findings, no items will be excluded at this stage to allow a comprehensive item set to be evaluated via an EFA in a larger sample (n ≈ 240) (36). This approach aims to ensure that potential factor structures are not prematurely constrained, allowing for the empirical determination of item groupings and loadings.

The group comparison showed statistically significant differences for three items, two of high relevance (nervousness and irritability) and one of lower relevance (distraction). These results correspond to those of the worst itch intensity and quality of life of the respective group and support the inclusion of all items for a valid item reduction with a larger sample.

This cross-sectional, pilot study was the first step in developing a pruritus-associated stress scale for CP. Several of the generated scale items achieved an impact score above the defined suitability limit (2.3). Future phases of the study will focus on further item reduction and the validation of the PASS.

## Data Availability

All data produced in the present work are contained in the manuscript.

https://bmjopen.bmj.com/content/12/1/e057596

## DATA AVAILABILITY STATEMENT

The authors confirm that the data supporting the findings of this study are available within the article and its supplementary materials.

## ETHICS DECLARATIONS & TRIAL REGISTRY INFORMATION

### IRB approval status

Reviewed and approved by the Ethics Committee of the State Medical Association of Westfalen-Lippe, Münster, Germany; approval # 2024-884-f-S.

## REFERENCES

1. Ständer S, Zeidler C, Augustin M, Darsow U, Kremer AE, Legat FJ, et al. S2k guideline: Diagnosis and treatment of chronic pruritus. J Dtsch Dermatol Ges 2022; 20: 1387–1402.

2. Ständer S, Schmelz M, Lerner E, Murota H, Nattkemper L, Reich A, et al. A multidisciplinary Delphi consensus on the modern definition of pruritus: Sensation and disease. J Eur Acad Dermatol Venereol. 2025

3. Ständer S. Classification of Itch. Curr Probl Dermatol 2016; 50: 1–4.

4. Ficheux A-S, Brenaut E, Schut C, Dalgard FJ, Bewley A, van Middendorp H, et al. Predictors of perceived stress, perceived stigmatization, and body dysmorphia in patients with chronic prurigo/prurigo nodularis: Results from an observational cross-sectional multicenter European study in 17 countries. J Am Acad Dermatol 2025; 92: 1056–1063.

5. Zeidler C, Kupfer J, Dalgard FJ, Bewley A, Evers AWM, Gieler U, et al. Dermatological patients with itch report more stress, stigmatization experience, anxiety and depression compared to patients without itch: Results from a European multi-centre study. J Eur Acad Dermatol Venereol 2024; 38: 1649–1661.

6. Misery L, Chesnais M, Merhand S, Aubert R, Bru MF, Legrand C, et al. Perceived stress in four inflammatory skin diseases: an analysis of data taken from 7273 adult subjects with acne, atopic dermatitis, psoriasis or hidradenitis suppurativa. J Eur Acad Dermatol Venereol 2022; 36: e623–e626.

7. Mochizuki H, Lavery MJ, Nattkemper LA, Albornoz C, Valdes Rodriguez R, Stull C, et al. Impact of acute stress on itch sensation and scratching behaviour in patients with atopic dermatitis and healthy controls. Br J Dermatol 2019; 180: 821–827.

8. Mochizuki H, Schut C, Shevchenko A, Valdes-Rodriguez R, Nattkemper LA, Yosipovitch G. A Negative Association of Hypothalamic Volume and Perceived Stress in Patients with Atopic Dermatitis. Acta Derm Venereol 2020; 100: adv00129.

9. Buske-Kirschbaum A, Ebrecht M, Hellhammer DH. Blunted HPA axis responsiveness to stress in atopic patients is associated with the acuity and severeness of allergic inflammation. Brain Behav Immun 2010; 24: 1347–1353.

10. Suárez AL, Feramisco JD, Koo J, Steinhoff M. Psychoneuroimmunology of psychological stress and atopic dermatitis: pathophysiologic and therapeutic updates. Acta Derm Venereol 2012; 92: 7–15.

11. Ständer S. How acute stress impacts the itch-scratch cycle in atopic dermatitis: a clinical lesson. Br J Dermatol 2019; 180: 689–690.

12. Schut C, Weik U, Tews N, Gieler U, Deinzer R, Kupfer J. Coping as mediator of the relationship between stress and itch in patients with atopic dermatitis: a regression and mediation analysis. Exp Dermatol 2015; 24: 148–150.

13. Grandgeorge M, Misery L. Mediators of the relationship between stress and itch. Exp Dermatol 2015; 24: 334–335.

14. Toussaint A, Weigel A, Löwe B. The overlooked burden of persistent physical symptoms: a call for action in European healthcare. Lancet Reg Health Eur 2025; 48: 101140.

15. Reis D, Lehr D, Heber E, Ebert DD. The German Version of the Perceived Stress Scale (PSS-10): Evaluation of Dimensionality, Validity, and Measurement Invariance With Exploratory and Confirmatory Bifactor Modeling. Assessment 2019; 26: 1246–1259.

16. Kocalevent R-D, Levenstein S, Fliege H, Schmid G, Hinz A, Brähler E, et al. Contribution to the construct validity of the Perceived Stress Questionnaire from a population-based survey. J Psychosom Res 2007; 63: 71–81.

17. Finlay AY, Khan GK. Dermatology Life Quality Index (DLQI)--a simple practical measure for routine clinical use. Clin Exp Dermatol 1994; 19: 210–216.

18. Desai NS, Poindexter GB, Monthrope YM, Bendeck SE, Swerlick RA, Chen SC. A pilot quality-of-life instrument for pruritus. J Am Acad Dermatol 2008; 59: 234–244.

19. Krause K, Kessler B, Weller K, Veidt J, Chen SC, Martus P, et al. German version of ItchyQoL: validation and initial clinical findings. Acta Derm Venereol 2013; 93: 562–568.

20. Zigmond AS, Snaith RP. The hospital anxiety and depression scale. Acta Psychiatr Scand 1983; 67: 361–370.

21. Ständer S, Augustin M, Reich A, Blome C, Ebata T, Phan NQ, et al. Pruritus assessment in clinical trials: consensus recommendations from the International Forum for the Study of Itch (IFSI) Special Interest Group Scoring Itch in Clinical Trials. Acta Derm Venereol 2013; 93: 509–514.

22. Schober P, Boer C, Schwarte LA. Correlation Coefficients: Appropriate Use and Interpretation. Anesth Analg 2018; 126: 1763–1768.

23. Weller K, Donoso T, Magerl M, Aygören-Pürsün E, Staubach P, Martinez-Saguer I, et al. Validation of the Angioedema Control Test (AECT)-A Patient-Reported Outcome Instrument for Assessing Angioedema Control. J Allergy Clin Immunol Pract 2020; 8: 2050–2057.e4.

24. Tavakol M, Dennick R. Making sense of Cronbach’s alpha. Int J Med Educ 2011; 2: 53– 55.

25. Kim HJ, Park JB, Lee JH, Kim I-H. How stress triggers itch: a preliminary study of the mechanism of stress-induced pruritus using fMRI. Int J Dermatol 2016; 55: 434–442.

26. Kim B, Rothenberg ME, Sun X, Bachert C, Artis D, Zaheer R, et al. Neuroimmune interplay during type 2 inflammation: Symptoms, mechanisms, and therapeutic targets in atopic diseases. J Allergy Clin Immunol 2024; 153: 879–893.

27. Dinhof C, Humer E, Haider K, Rabenstein R, Jesser A, Pieh C, et al. Comprehensive examination of support needs and mental well-being: a mixed-method study of the Austrian general population in times of crisis. Front Public Health 2024; 12: 1345796.

28. Ferreira BR, Misery L. Psychopathology Associated with Chronic Pruritus: A Systematic Review. Acta Derm Venereol 2023; 103: adv8488.

29. Woodhead E, Cronkite R, Finlay A, Wong J, Haverfield M, Timko C. The role of depression course on life functioning and coping outcomes from baseline through 23-year follow-up. J Ment Health 2022; 31: 348–356.

30. Lugović-Mihić L, Meštrović-Štefekov J, Pondeljak N, Dasović M, Tomljenović-Veselski M, Cvitanović H. Psychological Stress and Atopic Dermatitis Severity Following the COVID-19 Pandemic and an Earthquake. Psychiatr Danub 2021; 33: 393–401.

31. Pustišek N, Šitum M, Vurnek Živković M, Ljubojević Hadžavdić S, Vurnek M, Niseteo T. The significance of structured parental educational intervention on childhood atopic dermatitis: a randomized controlled trial. J Eur Acad Dermatol Venereol 2016; 30: 806– 812.

32. Meštrović-Štefekov J, Lugović-Mihić L, Hanžek M, Bešlić I, Japundžić I, Karlović D. Salivary Cortisol Values and Personality Features of Atopic Dermatitis Patients: A Prospective Study. Dermatitis 2022; 33: 341–348.

33. Kobusiewicz AK, Tarkowski B, Kaszuba A, Lesiak A, Narbutt J, Zalewska-Janowska A. The relationship between atopic dermatitis and atopic itch in children and the psychosocial functioning of their mothers: A cross-sectional study. Front Med (Lausanne) 2023; 10: 1066495.

34. Lind N, Nordin M, Palmquist E, Nordin S. Psychological distress in asthma and allergy: the Västerbotten Environmental Health Study. Psychol Health Med 2014; 19: 316–323.

35. Balieva F, Schut C, Kupfer J, Lien L, Misery L, Sampogna F, et al. Perceived stress in patients with inflammatory and non-inflammatory skin conditions. An observational controlled study among 255 Norwegian dermatological outpatients. Skin Health Dis 2022; 2: e162.

36. Costello AB, Osborne J. (2005) Best practices in exploratory factor analysis: four recommendations for getting the most from your analysis. Practical Assessment, Research, and Evaluation: Vol. 10, Article 7.

