## Supplemental Table SI for "Development of the Pruritus-Associated Stress Scale: A Cross-Sectional Pilot Study in Adults with Atopic Dermatitis, Chronic Prurigo and Chronic Pruritus on Non-Lesional Skin"

**SUPPLEMENT**

**Table SI: Interview questions to assess pruritus-associated stress within the item generation phase**

| Question No. | Question |
|---|---|
| **1** | In the past month, have you felt nervous or stressed due to pruritus? If so, how does your pruritus-associated stress manifest itself? |
| **2** | How frequently have you felt nervous or stressed due to pruritus in the past month? |
| **3** | Can you describe specific situations in which pruritus and stress are directly related for you? |
| **4** | In the past month, have you experienced difficulties falling asleep or staying asleep as a result of pruritus-associated stress? If yes, please describe these difficulties in more detail and indicate how often they occurred. |
| **5** | Do you have any strategies or methods for coping with stress triggered by pruritus? |
| **6** | Do you notice a stronger or more frequent urge to scratch when you are stressed? Do you ever scratch in response to stress even if you are not experiencing pruritus? (If yes, how often does this occur?) |
